# Saliva as a Reliable and Non-Invasive Sample for Detecting Influenza A in Severe Acute Respiratory Infection Cases

**DOI:** 10.1101/2025.05.20.25326848

**Authors:** Junko S. Takeuchi, Nobuaki Matsunaga, Ai Tsukada, Noriko Iwamoto, Noriko Fuwa, Takahiro Ichikawa, Yasuyuki Kato, Yuka Tomita, Hiroki Kitagawa, Masaya Yamato, Tetsuji Aoyagi, Hideharu Hagiya, Ryota Hase, Shuji Hatakeyama, Tohru Inaba, Koichi Izumikawa, Yoshio Takesue, Moto Kimura, Norio Ohmagari

**Author notes:** Corresponding Author: Nobuaki Matsunaga, MD, MPH, PhD, AMR Clinical Reference Center, Japan Institute for Health Security. 1-21-1 Toyama, Shinjuku-ku, Tokyo, 162-8655, JAPAN. (JST); (MK); (NM); (AT); (NI); (NF); (NO); (Takahiro I); (YK); (Yuka T); (HK); (MY); (TA); (HH); (RH); (SH); (Tohru I); (KI); (Yoshio T).

## Abstract

Nasopharyngeal swab sampling remains the gold standard for influenza A diagnosis but has limitations, such as dependence on medical staff, invasiveness, nosocomial transmission, and occupational exposure risk. This study aimed to investigate whether saliva and nasal vestibular swabs are suitable non-invasive alternatives to nasopharyngeal swabs for influenza A detection in severe acute respiratory infection (SARI) cases. Paired saliva and nasal vestibular swabs were collected on the same day from 16 cases diagnosed with influenza. Saliva samples demonstrated a higher sensitivity (87.5%) than did nasal vestibular swabs (31.3%) in RT-qPCR, when compared with the diagnostic results obtained from nasopharyngeal swabs. While nasal vestibular swabs showed inconsistent results, saliva samples consistently tested positive, particularly within 7 days of symptom onset (100% positive agreement). In addition, diagnosis with RT-qPCR is often delayed because it requires trained laboratory technicians and facilities with appropriate laboratory settings. Therefore, the GenPad®, a rapid diagnostic device, was evaluated using saliva samples and showed promising performance (92.9%) compared with the efficiency of RT-qPCR. Factors such as the location of infection (upper vs. lower respiratory tract infections), sample collection timing, pre-collection instructions, and nucleic acid extraction possibly contributed to the detection efficiency. Despite the small sample size and lack of influenza-negative controls, our findings support saliva as a viable self-collected sample for influenza A diagnosis and surveillance programs. Non-invasive sampling mitigates discomfort, minimizes infection risk for healthcare workers, and improves testing capacity, particularly under frequent staff shortages during pandemics.

## 1. Introduction

The Coronavirus Disease 2019 (COVID-19) pandemic has prompted a more comprehensive surveillance of acute respiratory infections (ARIs), including COVID-19, Respiratory Syncytial Virus (RSV) infection, and influenza, worldwide [1]. Countries use surveillance systems such as the Respiratory Virus Hospitalization Surveillance Network (RESP-NET; the United States) [2], the Severe Acute Respiratory Infection Watch (SARI Watch; the United Kingdom) [3], and the European Respiratory Virus Surveillance Summary (ERVISS; European Union) [4]. In Japan, clinical information and specimens derived from severe acute respiratory infections (SARIs) have been added to the Infectious Disease Clinical Research Network With National Repository (iCROWN, the successor to REBIND) since September 2024 [5]. Furthermore, on April 7, 2025, ARI has been designated a “Category V Infectious Diseases” under Japan’s Infectious Disease Control Law [6] and is subjected to sentinel surveillance.

As a representative SARI/ARI, influenza requires early diagnosis to appropriately guide antiviral therapy and patient isolation for effective disease control [7,8]. Antiviral treatment is most effective when initiated within 48 h of symptom onset, helping to reduce the likelihood of hospitalization or the need for critical care, as well as to prevent further transmission within the community. However, at the beginning of the COVID-19 pandemic, two major challenges in diagnosing severe acute respiratory syndrome coronavirus 2 (SARS-CoV-2) were highlighted. First, sample collection relied exclusively on healthcare workers. Second, reverse transcription real-time quantitative PCR (RT-qPCR)-based diagnosis was delayed due to the need for specialized personnel and facilities. In response, self-sampling and point-of-care testing (POCT) methods for molecular diagnosis were developed [9].

Although nasopharyngeal swab sampling remains the gold standard for influenza diagnosis, it requires medical personnel and can be uncomfortable, especially for children. Amid pandemics, staff shortages and the risk of aerosol generation during nasopharyngeal swab sampling further complicate this approach. Less-invasive, self-collectable methods such as saliva or nasal vestibular (anterior nasal cavity) swab sampling offer practical alternatives. These simpler collection methods may also increase participation rates in clinical studies and surveillance programs. Thanks to the high sensitivity of RT-qPCR, detection rates across different sample types in influenza virus may be comparable [10]. Multiple studies on influenza detection have reported high concordance rates (93%–100%) between saliva and nasopharyngeal samples [11–18]. However, studies comparing saliva and anterior nasal swab samples remain limited. Although two studies have reported that saliva appeared less effective than self-collected anterior nasal swabs for detecting influenza A, these studies included only a small number of influenza-positive cases (n=8 [19] and n=1 [20], respectively). Therefore, the evidence remains insufficient to draw firm conclusions about differences in detection performance between saliva and anterior nasal samples.

RT-qPCR is the gold standard for influenza detection because of its high sensitivity. However, it typically takes 2–3 h, including RNA extraction, and requires specialized facilities and a stable power supply, limiting its application in smaller or rural clinical setups and in urgent settings [21]. To address these challenges, the GenPad® (MIRAI GENOMICS, Kanagawa, Japan) represents a promising alternative. It utilizes isothermal nucleic acid amplification based on SmartAmp (a Smart Amplification Process) and fluorescence detection [22,23], offering a portable, battery-operated solution with results within 50 min. However, few studies have examined its performance using clinical samples.

This study therefore addresses the two key questions in SARI diagnostics: (i) can saliva or nasal vestibular swab samples serve as suitable alternatives to nasopharyngeal swab samples, and (ii) can the GenPad® provide a reliable option for influenza using saliva samples.

## 2. Material and methods

### 2.1. Ethics statement

This study was approved by the Ethics Review Committee of the National Center for Global Health and Medicine, Japan (NCGM-S-004948). All methods were carried out in accordance with relevant guidelines and regulations.

### 2.2. Study participants

We recruited inpatients who met certain criteria at the time of admission in Japan. The case-cohort criteria for SARI were:

i. All of A−C: (A) Respiratory pathogens were detected or strongly suspected. (B) Airway symptoms, such as cough. (C) At the time of admission, the patient was not in a care facility, undergoing tracheotomy, or having confirmed significant dysphagia.
ii. One or more of D−F: (D) Chest imaging showed infiltrates. (E) SpO2 was ≤94% under room air conditions. (F) New oxygen therapy, invasive mechanical ventilation, or non-invasive mechanical ventilation were introduced.

All patients gave informed consent for participation in the prospective observation study.

Sixteen patients classified as having SARIs and diagnosed with influenza participated in this study. Patients were diagnosed at day 0−4 (median: 1) after onset at each medical facility using one of the following methods with nasopharyngeal swab samples: Film Array test, Rapid Test kit, or antigen test (the day of onset was designated as day 0). There were 13 (81.25%) male participants aged 16–82 (median: 67.5) years. All clinical samples were collected 1−9 (median: 3.5) days after symptom onset. Paired samples (saliva and nasal vestibular samples) were collected from each patient on the same day. The nasal vestibular samples were collected using a swab and preserved in 3 mL of Universal Transport Medium (FLOQSwabs® and UTM®, 307C;

Copan, Brescia, Italy). Saliva samples were collected in sterile tubes without preservative medium using a saliva collector (SP1-RO0003-56, AXIS Inc., Ibaraki, Japan). The participants avoided gargling, brushing their teeth, or eating within 30 min before sample collection. All residual samples were stored at −20 or −80 °C until use.

### 2.3. Nucleic acid extraction

Each saliva sample (150 µL) was diluted with an equal volume of phosphate-buffered saline without calcium and magnesium [PBS(-)], vigorously mixed, and spun-down. Nucleic acid (60 µL) was extracted from 200 µL of sample, either saliva [diluted two-fold with PBS(-)] or nasal vestibular swab suspended in 3 mL of UTM, using a KingFisher APEX System (Thermo Fisher Scientific, Waltham, MA, USA) and the MagMAX Prime Viral/Pathogen NA Isolation Kit (A58145; Thermo Fisher Scientific), with 60% ethanol used as the second wash buffer.

### 2.4. Reverse transcription real-time quantitative polymerase chain reaction

RT-qPCR was performed according to the Influenza Diagnostic Manual (5th Edition, August 2023) published by the National Institute of Infectious Diseases (NIID), Japan [24]. Briefly, RT-qPCR was performed on a QuantStudio®3 real-time PCR system (Thermo Fisher Scientific) using AgPath-IDTM One-Step RT-PCR Reagents (AM1005; Thermo Fisher Scientific) with the following primer/probe sets: the influenza A set comprised [MP-39-67For (5’-CCMAGGTCGAAACGTAYGTTCTCTCTATC-3’), MP-183-153Rev (5’-TGACAGRATYGGTCTTGTCTTTAGCCAYTCCA-3’), and MP-96-75ProbeAs (5’-FAM-ATYTCGGCTTTGAGGGGGCCTG-MGB-3’)]; the influenza B set consisted of [NIID-TypeB TMPrimer-F1 (5’-GGAGCAACCAATGCCAC-3’, NIID-TypeB TMPrimer-R1 (5’-GTKTAGGCGGTCTTGACCAG-3’), and NIID-TypeB Probe2 (5’-FAM-ATAAACTTYGAAGCAGGAAT-MGB-3’)]. Subsequently, 5 µL of the extracted nucleic acid was used as the template in 25 µL of an RT-qPCR reaction mixture using the following conditions: 50 °C for 10 min, 95 °C for 10 min, followed by 45 cycles of 95 °C for 15 sec, 56 °C for 30 sec, and 72 °C for 15 sec. The data was analyzed using QuantStudio^TM^ Design & Analysis Software v1.5.2. The test was considered successful when amplification and no-amplification curves were confirmed in the positive and negative (nuclease-free water) controls, respectively. A sample was considered positive when the amplification curve increased. The positive RNA controls were kindly provided by the NIID [for influenza A: (A/Wisconsin/67/2022, AH1 pdm S2 +hCK1); for influenza B: (B/Austria/1359417/2021, B Victoria S1C4/ C2 +HCK1)].

### 2.5. GenPad system

The GenPad® Smart CoV2/FluA/FluB Research Use Only kit (RR072; MIRAI GENOMICS) was used on a GenPad® device according to the manufacturer’s protocol with minor modifications. In contrast to using a swab-collected saliva sample, as instructed in the kit, 150 µL of saliva sample was used. Briefly, a saliva sample (150 µL) was added to a tube containing a swab suspension buffer (SSB; 2.0 mL). After inversion mixing, a nozzle cap was attached to the tube, and the extracted mixture was added to the Sample Loading Level of the GenPad Smart test cartridge. When the filter of the nozzle cap became clogged due to the viscosity of saliva sample, it was replaced with a new cap.

### 2.6. Statistical analysis

Comparisons of RT-qPCR results between saliva and nasal vestibule samples, as well as between RT-qPCR and GenPad results using saliva samples, were evaluated using exact McNemar test (`mcnemar.exact`) from the *exact2x2* package (v1.7.0) [25]. Positive and negative agreement rates were obtained using the `epi.tests` function of the *epiR* package (v2.0.88), with 95% confidence intervals (CIs) calculated using the Wilson method [26]. To assess differences in Threshold Cycle (Ct) values between saliva and nasal vestibular swabs, survival analysis was performed with undetected samples treated as right-censored at Ct = 40. Kaplan–Meier curves were generated using the `survfit` function to compare the proportion of undetected results between sample types, and differences were evaluated using a stratified log-rank test with the `survdiff` function (stratified by case No.) from the *survival* package (v3.8-3). The correlation and *p*-value between RT-qPCR Ct values and GenPad® Threshold Time (Tt) values were calculated using Spearman’s rank correlation. All statistical analyses were performed in R 4.3.1 (R Foundation for statistical Computing, Vienna, Austria).

## 3. Results

We tested paired saliva and nasal vestibular samples collected on the same day from 16 patients classified as having SARI and diagnosed with influenza between December 2024 and March 2025 in Japan. All patients, except for case No. 8, had either no history of influenza vaccination or an unknown vaccination status. All samples that tested positive by RT-qPCR were identified as influenza A, and no influenza B was detected. Although the RT-qPCR in this study was qualitative and lacked a calibration curve, all samples were tested on the same plate, except for No. 16, allowing Ct-based comparisons. Although No. 16 was tested separately, its positive control Ct value (31.4) was equivalent to the other assay (31.6), so it was included in the analysis.

### 3.1. Comparison with diagnostic results

We first compared the diagnostic results obtained from nasopharyngeal swabs (collected at medical facilities using one of the following methods: Film Array test, Rapid Test kit, or antigen test) with the RT-qPCR results of the saliva and nasal vestibular samples. As this study included only cases diagnosed with influenza, a positive agreement rate could be calculated; 87.5% (14 of 16) for saliva and 31.3% (5 of 16) for nasal vestibular swab samples. Diagnosis using nasopharyngeal swabs was performed 0–4 days (median: 1) after symptom onset, which may have led to an underestimation of the sensitivity of saliva and nasal vestibular samples that were collected 1−9 days (median: 3.5) after symptom onset. This temporal discrepancy in sampling should be considered when interpreting the positive agreement rates.

### 3.2. Saliva versus nasal vestibular samples

Subsequently, we compared the RT-qPCR results of the paired saliva and nasal vestibular samples (Figure 1). The positive, negative, and total agreement rates were 35.7% (95% CI: 16.3%–61.2%), 100% (34.2%–100%), and 43.8% (23.1%–66.8%), respectively, using saliva as the reference (Table 1). A significant difference in detection sensitivity between saliva and nasal vestibular samples was observed using McNemar’s exact test (p=0.0039), indicating that the positivity rate was higher in saliva samples compared with nasal vestibular samples. When saliva Ct values were ≥37, those of nasal vestibular samples were undetectable (n=6), whereas nasal vestibular samples were also detectable when saliva Ct values were ≤30 (n=3) (Figure 1A). Furthermore, differences in Ct values between the two sample types were evaluated using survival analysis, treating undetected samples as right-censored at a Ct value of 40. Kaplan–Meier curves demonstrated that the proportion of undetected samples differed between the two sample types, and a stratified log-rank test (stratified by case No.) confirmed that this difference was statistically significant (p=0.0075) (Figure 1B).

**Figure 1.**
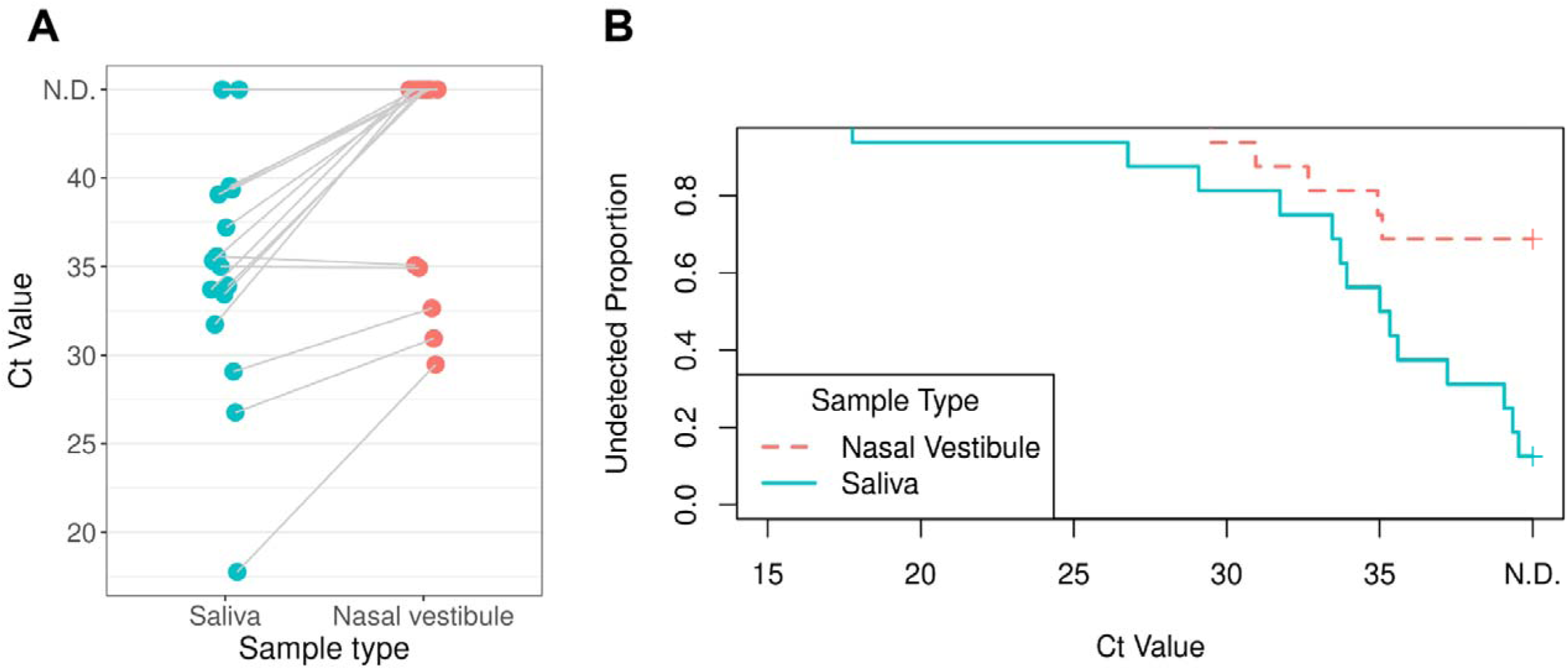
Ct values obtained by reverse transcription quantitative PCR (RT-qPCR) among different sample types. (A) Comparison of Ct values obtained by reverse RT-qPCR among different sample types. (B) Comparison of the proportion of undetected samples between saliva and nasal vestibule samples using Kaplan–Meier analysis. Undetected samples were treated as right-censored at Ct=40. Blue and pink circles/lines represent saliva and nasal vestibular samples, respectively. N.D.; not detected (no detectable amplification).

**Table 1.** Comparison of RT-qPCR using saliva and nasal vestibule samples.

| RT-qPCR | Nasal vestibule |  |  |  |
| --- | --- | --- | --- | --- |
|  |  | Positive | Negative | Total |
| Saliva | Positive | 5 | 9 | 14 |
|  | Negative | 0 | 2 | 2 |
|  | **Total** | **5** | **11** | **16** |

### 3.3. Impact of antiviral treatment and sampling timing

Next, we evaluated the relationship between clinical information and the Ct values of the RT-qPCR. Prior to sample collection, antiviral treatment had been administered in 11 out of 16 cases: peramivir was given to Nos. 1–7 and 14; oseltamivir to Nos. 8, 13, and 15; and no antiviral treatment was given to Nos. 9–12 and 16.

Although Nos. 7 and 15 (both of whom had received antiviral treatment) tested negative under RT-qPCR in both saliva and nasal vestibular swab samples, the fact that other treated patients yielded positive results suggested that antiviral treatment alone does not fully account for the variability in detection sensitivity. In contrast, the number of days from onset to sample collection appears to be more relevant: Nos. 7 and 15 were collected on day 8 or 9 post-onset, when viral loads had likely been reduced, possibly explaining the negative results (Figure 2).

**Figure 2.**
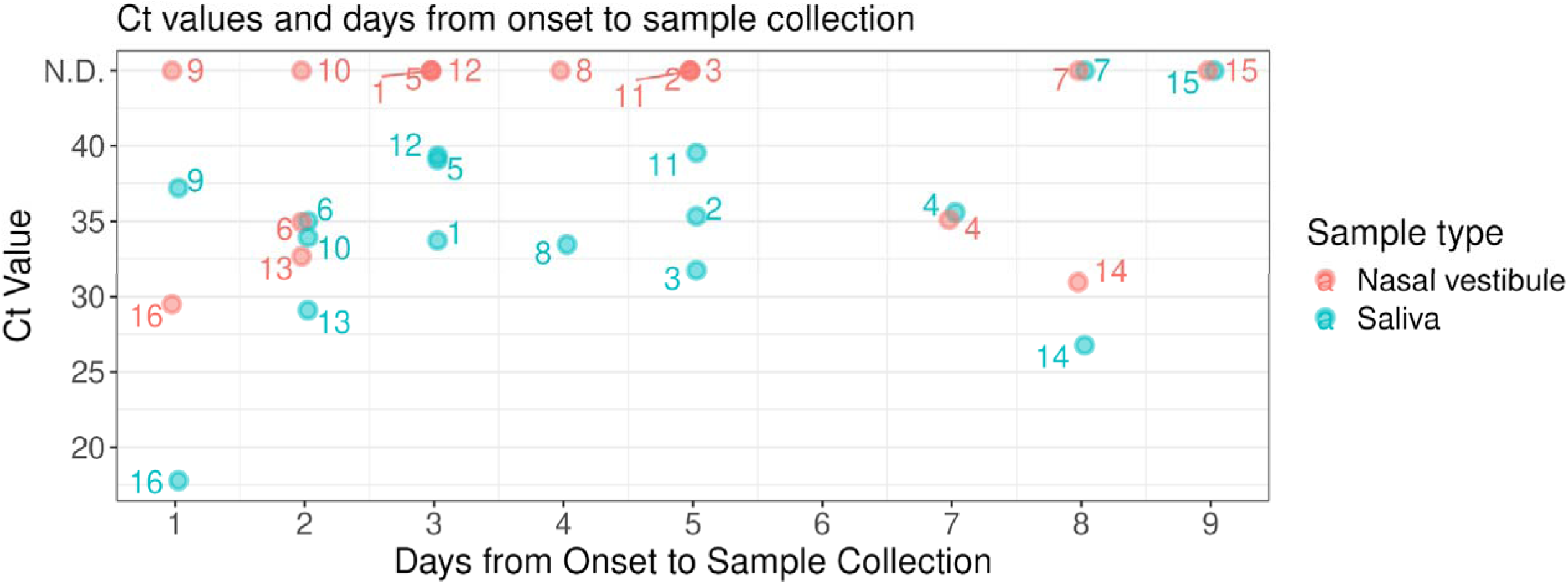
Comparison of RT-qPCR Ct values with time from symptom onset to sample collection. Days from onset to sample collection are shown on the x-axis, and Ct values are on the y-axis. Blue and pink circles represent saliva and nasal vestibular samples, respectively.

Sample No. 14 was collected on day 8 and had the second-lowest Ct value (second-highest viral load). However, this patient was immunosuppressed due to complications following hematopoietic stem cell transplantation and was receiving multiple immunosuppressive therapies, suggesting prolonged viral persistence.

### 3.4. GenPad® rapid test performance

Using saliva samples, we compared the GenPad® and RT-qPCR (Figure 3). The positive, negative, and total agreement rates were 92.9% (68.5%–98.7%), 100% (34.2%–100%), and 93.8 (71.7%–98.9%), respectively (Table 2). No significant differences in detection sensitivity between the two tests were observed using McNemar’s exact test (p=1). One discordant sample No. 12 yielded a Ct value (39.5) using RT-qPCR but was not detected by the GenPad®, suggesting that the viral load was near or below the detection limit of the GenPad®. Although Spearman’s rank correlation coefficient between Tt/Ct values of the GenPad® and RT-qPCR was moderately high (r=0.522), it did not reach statistical significance (p=0.067). The GenPad® Smart CoV2/FluA/FluB kit detects Influenza A, B and SARS-CoV-2; however, only influenza A was detected in this study, consistent with RT-qPCR (although the detection of SARS-CoV-2 was not included in the RT-qPCR).

**Figure 3.**
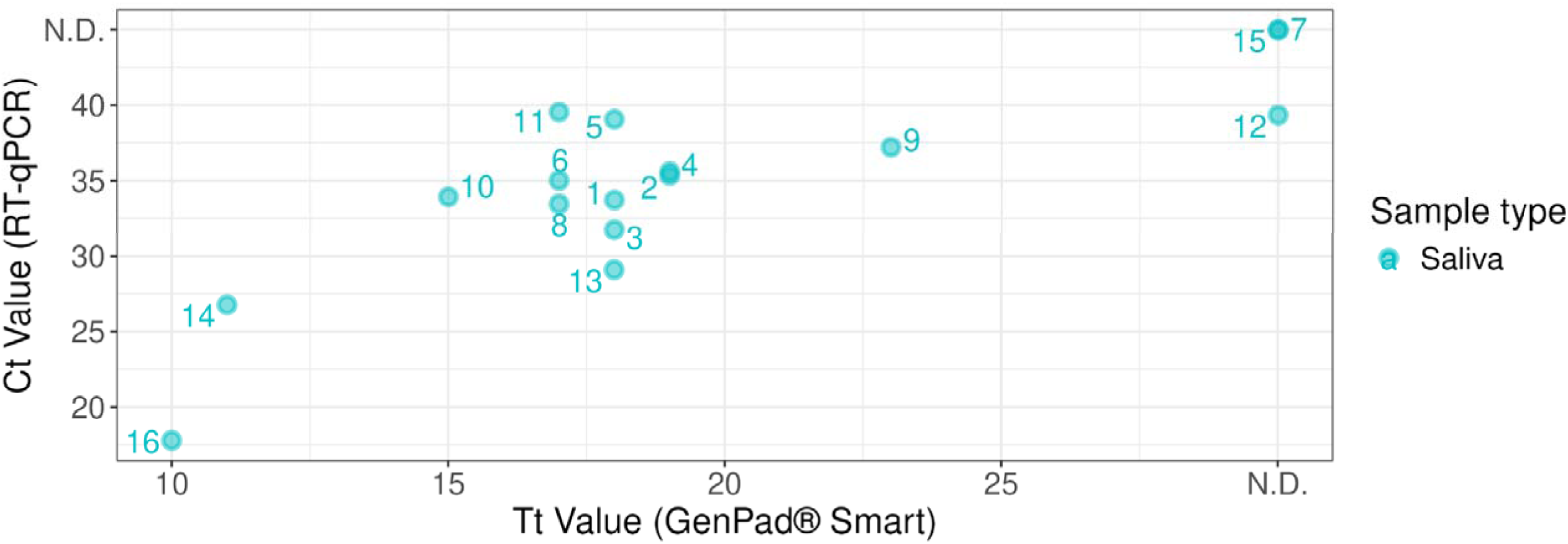
Correlation of Tt/Ct values between the GenPad and RTqPCR methods. Blue circles indicate Tt/Ct values of saliva samples, respectively. N.D.; not detected (no detectable amplification).

**Table 2.** Comparison of RT-qPCR and GenPad using saliva sample.

| Saliva | GenPad |  |  |  |
| --- | --- | --- | --- | --- |
|  |  | Positive | Negative | Total |
| RT-qPCR | Positive | 13 | 1 | 14 |
|  | Negative | 0 | 2 | 2 |
|  | **Total** | **13** | **3** | **16** |

## 4. Discussion

This study demonstrates that saliva is a more reliable non-invasive sample for influenza A detection than nasal vestibular swabs in SARI cases, particularly within 7 days post-onset. Our findings differ from those of Goff et al., which demonstrated lower sensitivity of saliva compared to self-collected anterior nasal swabs [19]; however, their study included only 8 influenza-positive cases and statistical significance was not evaluated. Nevertheless, several factors may explain the apparent discrepancy between the two studies. First, our SARI cases likely included viral pneumonia (lower respiratory tract infections [LRTIs]), whereas, Goff et al. [19] reported non-severe seasonal influenza cases (upper respiratory tract infections [URTIs]). These clinical differences may have involved differences in viral load distribution within the respiratory tract, potentially affecting the sensitivity of sample types. Second, our study followed strict pre-collection conditions, asking the participants to avoid gargling, brushing their teeth, or eating within 30 min before sample collection based on the guidelines of the Ministry of Health, Labour and Welfare. These actions interfere with viral detection in saliva samples, according to studies on SARS-CoV-2 [27,28], possibly because the impurities in saliva samples may affect PCR efficiency [29]. In contrast, in the study by Goff et al. [19] conducted before the COVID-19 pandemic, these conditions were not established for saliva collection, which may have affected virus detection. However, even when similar pre-collection conditions are applied, other factors may still influence detection sensitivity. For example, another study by Norizuki et al. [30], who tested asymptomatic travelers for SARS-CoV-2 and implemented similar pre-collection conditions for saliva sampling, reported low detection rates in saliva samples. Norizuki et al. [30] noted that the participants self-collected samples without supervision at busy airport quarantine stations, which may have affected their results. Thus, supervised saliva collection under conditions that meet pre-collection requirements may be preferable. Next, we extracted nucleic acids from the saliva samples to effectively remove impurities, ensuring that their impact on the results was minimized. In contrast, Goff et al. [19] used several rapid diagnostic kits, a condition where the substances in the saliva might have interfered with the outcomes.

The lower and variable sensitivity of nasal vestibular swabs may be related to the location of respiratory symptoms (presence of nasal discharge, URTIs vs. LRTIs), age group (children or adults), sampling techniques (rubbed vigorously or lightly, swab suspension ratio), or collection area (anterior or posterior of the nasal cavity). These results indicate that nasal vestibular swab samples may pose a risk of sampling bias. Additionally, Branche et al. reported that RT-qPCR revealed higher mean viral loads in sputum than in nasal or throat swabs for influenza A [31]. Therefore, sputum should also be considered as a representative sample for influenza detection.

Most healthy volunteers ceased shedding influenza virus by day 6 or 7 [32,33], though shedding may last longer in children or immunocompromised patients [34,35]. In our study, all saliva samples collected at ≤7 days of onset tested positive, with saliva-based RT-qPCR showing 100% positive agreement with nasopharyngeal diagnostics (Figure 2). Although pediatric cases were not included in this study, the only minor case (aged 15−19, No. 4) tested positive in both saliva and nasal vestibular samples on day 7. Furthermore, although it is just one case, we confirmed that an immunosuppressed patient (No. 14) can retain higher levels of the virus for an extended period, as previously reported [34].

A saliva-based rapid diagnostic kit, the GenPad®, demonstrated relatively good performance (positive agreement rate 92.9%). This aligns with a previous report showing a 100% positive concordance rate compared with that of the RT-qPCR method; however, the result was based on SARS-CoV-2 detection in saliva samples [36]. Suspension of the saliva samples in SSB containing ethanol and a chaotropic agent (guanidine thiocyanate), followed by filtration through a glass filter, may effectively contribute to the removal of impurities and elution of nucleic acids. Notably, 150 µL of saliva sample was directly treated with SSB in this study despite the kit’s protocol specifying swab-collected saliva; therefore, further evaluation is warranted. The COVID-19 pandemic highlighted the use of saliva as a sensitive and reliable diagnostic sample, especially for nucleic acid testing [18,37]. A growing body of evidence supports its use of saliva for detecting other respiratory pathogens including influenza. This may promote the approval of saliva-based diagnostic tests for influenza. Although not assessed in this study, saliva has reportedly shown lower sensitivity in immunochromatographic rapid antigen tests [38].

The strength of this study is the direct comparison of two non-invasive sample types using RT-qPCR and access to detailed clinical data, such as the treatment history and the symptom onset date. However, this study had some limitations. First, we did not evaluate nasopharyngeal swabs simultaneously. Second, the samples were collected 1 to 9 days after the onset of symptoms, and antiviral therapy had been initiated in 11 out of 16 cases. These factors (a prolonged interval between onset and testing or prior treatment) may have reduced viral loads in both saliva and nasal vestibular swab samples, potentially lowering overall detection sensitivity. Third, the samples were small-scale (n=16) and biased toward those obtained in SARI cases in Japan, preventing the generalizability of the results to a larger population or other settings. Fourth, because we did not include a control group of influenza-negative patients, we could not evaluate the negative agreement rate. Finally, while we confirmed the virus detection from saliva samples, further evaluation of viral characteristics are warranted, particularly to assess whether viable viruses can be isolated from saliva, as described in SARS-CoV-2 studies [39,40].

We demonstrated that the use of saliva, but not nasal vestibular swabs, is a potential non-invasive alternative to nasopharyngeal swabs for influenza A virus detection particularly within 7 days of symptom onset. Additionally, the GenPad® provides comparable results to RT-qPCR. These methods may reduce the burden on patients and healthcare settings, mitigate nosocomial infections, and increase participation rates when applied for clinical studies and surveillance purposes.

## Data Availability

The data that support the findings of this study, excluding those subjects to privacy or ethical restrictions, are available from the corresponding author upon reasonable request.

## Acknowledgements

We are grateful to the patients who contributed to this study. We thank clinical staff of the National Center for Global Health and Medicine, Japan, Dr. Atsushi Nagasaka [4], Dr. Saho Takaya [5], Ms. Masako Shikami [5], Dr. Retsu Fujita [5], Dr. Hisashi Wakayama [6], Dr. Naohiko Murata [6], Dr. Hiroki Ohge [7], and Dr. Norifumi Shigemoto [7] for providing the clinical samples and the valuable suggestions on the manuscript; Dr. Chiho Kitamura [9], Dr. Asami Nakayama [9], Dr. Shinnosuke Fukushima [10], Dr. Kenta Nakamoto [10], Dr. Haruhiko Ishioka [12], Dr. Takaaki Nakaya [13], Mr. Yukiji Yamada [13], Dr. Hiroshi Ikai [13], Mr. Takeo Okashita [13], and Dr. Masato Tashiro [14] for insightful comment on the manuscript; Dr. Tadaki Suzuki [16] and Dr. Sho Miyamoto [16] for providing the positive RNA control; Dr. Nobuhiro Takemae [16] for valuable advice on the RT-qPCR protocol; Dr. Yosuke Shimizu [17] for guidance on statistical analysis; and Ms. Azusa Kamikawa [1], Dr. Sakino Takayanagi-Nishisako [1], Ms. Emiko Hatano [1], Dr. Kento Fukano [1], and Ms. Ryoko Tamura [1] for their experimental support. (The affiliation identifiers listed here are the same as those listed in the Author section. In addition, affiliations [16] and [17] correspond to the National Institute of Infectious Diseases, Tokyo, Japan, and Department of Data Sciences, Center for Clinical Sciences, Japan Institute for Health Security, Tokyo, Japan, respectively.)

## Author contributions

Conceptualization, Funding acquisition: NM.

Methodology: NM, NI, NF, Takahiro I, YK, Yuka T, HK, MY, TA, HH, RH, SH, Tohru I, KI, and Yoshio T.

Resources: NI, NF, Takahiro I, YK, Yuka T, HK, and MY.

Software, Formal analysis, Investigation, Visualization, Writing - Original Draft: JST.

Validation, Data Curation, Project administration: AT and NM.

Supervision: MK and NO.

Writing - Review & Editing: all authors

## References

1. Matsunaga N, Ohmagari N: Surveillance of Severe Acute Respiratory Infections (SARI) in Europe, the United States, and Japan. (Japanese). Infectious Agents Surveillance Report (IASR). 2024, 45:200-1

2. Respiratory Virus Hospitalization Surveillance Network (RESP-NET). Accessed: April 10, 2025: https://www.cdc.gov/resp-net/dashboard/

3. The severe acute respiratory infection (SARI) Watch surveillance system. Accessed: April 10, 2025: https://www.gov.uk/government/publications/sources-of-surveillance-data-for-influenza-covid-19-and-other-respiratory-viruses/data-quality-report-national-flu-and-covid-19-surveillance-report#secondary-care-surveillance-sari-watch

4. The European Respiratory Virus Surveillance Summary (ERVISS) Accessed: April 10, 2025: https://erviss.org/

5. iCROWN. Accessed: May 12, 2025: https://nwp.ncgm.go.jp/index.html

6. Act on the Prevention of Infectious Diseases and Medical Care for Patients with Infectious Diseases. Accessed: May 10, 2025: https://www.japaneselawtranslation.go.jp/ja/laws/view/4822

7. Louie JK, Acosta M, Jamieson DJ, Honein MA, California Pandemic Working G: Severe 2009 H1N1 influenza in pregnant and postpartum women in California. N Engl J Med. 2010, 362:27–35 10.1056/NEJMoa0910444

8. Glezen WP: Clinical practice. Prevention and treatment of seasonal influenza. N Engl J Med. 2008, 359:2579-85 10.1056/NEJMcp0807498

9. Jayamohan H, Lambert CJ, Sant HJ, et al.: SARS-CoV-2 pandemic: a review of molecular diagnostic tools including sample collection and commercial response with associated advantages and limitations. Anal Bioanal Chem. 2021, 413:49–71. 10.1007/s00216-020-02958-1

10. Spencer S, Thompson MG, Flannery B, Fry A: Comparison of Respiratory Specimen Collection Methods for Detection of Influenza Virus Infection by Reverse Transcription-PCR: a Literature Review. J Clin Microbiol. 2019, 57. 10.1128/JCM.00027-19

11. Bilder L, Machtei EE, Shenhar Y, Kra-Oz Z, Basis F: Salivary detection of H1N1 virus: a clinical feasibility investigation. J Dent Res. 2011, 90:1136–9. 10.1177/0022034511413283

12. Galar A, Catalan P, Vesperinas L, et al.: Use of Saliva Swab for Detection of Influenza Virus in Patients Admitted to an Emergency Department. Microbiol Spectr. 2021, 9:e0033621. 10.1128/Spectrum.00336-21

13. Kim YG, Yun SG, Kim MY, et al.: Comparison between Saliva and Nasopharyngeal Swab Specimens for Detection of Respiratory Viruses by Multiplex Reverse Transcription-PCR. J Clin Microbiol. 2017, 55:226–33. 10.1128/JCM.01704-16

14. Sueki A, Matsuda K, Yamaguchi A, et al.: Evaluation of saliva as diagnostic materials for influenza virus infection by PCR-based assays. Clin Chim Acta. 2016, 453:71–4. 10.1016/j.cca.2015.12.006

15. To KK, Lu L, Yip CC, et al.: Additional molecular testing of saliva specimens improves the detection of respiratory viruses. Emerg Microbes Infect. 2017, 6:e49. 10.1038/emi.2017.35

16. To KKW, Yip CCY, Lai CYW, et al.: Saliva as a diagnostic specimen for testing respiratory virus by a point-of-care molecular assay: a diagnostic validity study. Clin Microbiol Infect. 2019, 25:372–8. 10.1016/j.cmi.2018.06.009

17. Yoon J, Yun SG, Nam J, Choi SH, Lim CS: The use of saliva specimens for detection of influenza A and B viruses by rapid influenza diagnostic tests. J Virol Methods. 2017, 243:15–9. 10.1016/j.jviromet.2017.01.013

18. Laxton CS, Peno C, Hahn AM, Allicock OM, Perniciaro S, Wyllie AL: The potential of saliva as an accessible and sensitive sample type for the detection of respiratory pathogens and host immunity. Lancet Microbe. 2023, 4:e837–e50. 10.1016/S2666-5247(23)00135-0

19. Goff J, Rowe A, Brownstein JS, Chunara R: Surveillance of Acute Respiratory Infections Using Community-Submitted Symptoms and Specimens for Molecular Diagnostic Testing. PLoS Curr. 2015, 7. 10.1371/currents.outbreaks.0371243baa7f3810ba1279e30b96d3b6

20. Jung EJ, Lee SK, Shin SH, et al.: Comparison of Nasal Swabs, Nasopharyngeal Swabs, and Saliva Samples for the Detection of SARS-CoV-2 and other Respiratory Virus Infections. Ann Lab Med. 2023, 43:434-42. 10.3343/alm.2023.43.5.434

21. Soroka M, Wasowicz B, Rymaszewska A: Loop-Mediated Isothermal Amplification (LAMP): The Better Sibling of PCR? Cells. 2021, 10.10.3390/cells10081931

22. Asai N, Nakamura A, Sakanashi D, et al.: Comparative study of SmartAmp assay and reverse transcription-polymerase chain reaction by saliva specimen for the diagnosing COVID-19. J Infect Chemother. 2022, 28:120–3. 10.1016/j.jiac.2021.09.011

23. Nagasawa S, Mori A, Hirata Y, et al.: SmartAmp method can rapidly detect SARS-CoV-2 in dead bodies. Forensic Sci Int. 2022, 331:111168. 10.1016/j.forsciint.2021.111168

24. The Influenza Diagnostic Manual, 5th edition (August 2023). Accessed: https://id-info.jihs.go.jp/relevant/manual/010/influenza20230829.pdf

25. Fay MP: Two-sided Exact Tests and Matching Confidence Intervals for Discrete Data. R J. 2010, 2:53-8

26. Tools for the Analysis of Epidemiological Data. Accessed: https://CRAN.R-project.org/package=epiR

27. Meister TL, Bruggemann Y, Todt D, et al.: Virucidal Efficacy of Different Oral Rinses Against Severe Acute Respiratory Syndrome Coronavirus 2. J Infect Dis. 2020, 222:1289–92. 10.1093/infdis/jiaa471

28. Costa MM, Benoit N, Tissot-Dupont H, et al.: Mouth Washing Impaired SARS-CoV-2 Detection in Saliva. Diagnostics (Basel). 2021, 11. 10.3390/diagnostics11081509

29. Alaeddini R: Forensic implications of PCR inhibition--A review. Forensic Sci Int Genet. 2012, 6:297-305. 10.1016/j.fsigen.2011.08.006

30. Norizuki M, Hachiya M, Motohashi A, et al.: Effective screening strategies for detection of asymptomatic COVID-19 travelers at airport quarantine stations: Exploratory findings in Japan. Glob Health Med. 2021, 3:107–11. 10.35772/ghm.2020.01109

31. Branche AR, Walsh EE, Formica MA, Falsey AR: Detection of respiratory viruses in sputum from adults by use of automated multiplex PCR. J Clin Microbiol. 2014, 52:3590–6. 10.1128/JCM.01523-14

32. Aoki FY, Boivin G: Influenza virus shedding: excretion patterns and effects of antiviral treatment. J Clin Virol. 2009, 44:255–61. 10.1016/j.jcv.2009.01.010

33. Carrat F, Vergu E, Ferguson NM, et al.: Time lines of infection and disease in human influenza: a review of volunteer challenge studies. Am J Epidemiol. 2008, 167:775–85. 10.1093/aje/kwm375

34. Weinstock DM, Gubareva LV, Zuccotti G: Prolonged shedding of multidrug-resistant influenza A virus in an immunocompromised patient. N Engl J Med. 2003, 348:867–8. 10.1056/NEJM200302273480923

35. Sato M, Hosoya M, Kato K, Suzuki H: Viral shedding in children with influenza virus infections treated with neuraminidase inhibitors. Pediatr Infect Dis J. 2005, 24:931–2. 10.1097/01.inf.0000180976.81055.ce

36. Miyashita A, Kurosawa T, Yajima T, Yamazaki E: Performance Evaluation of SARS-CoV-2 RNA Measuring Device GenPad. J Jpn Soc Lab Med. 2023, 71:684–7

37. Oliveira Neto NF, Caixeta RAV, Zerbinati RM, et al.: The Emergence of Saliva as a Diagnostic and Prognostic Tool for Viral Infections. Viruses. 2024, 16. 10.3390/v16111759

38. Kok J, Blyth CC, Foo H, et al.: Comparison of a rapid antigen test with nucleic acid testing during cocirculation of pandemic influenza A/H1N1 2009 and seasonal influenza A/H3N2. J Clin Microbiol. 2010, 48:290–1. 10.1128/JCM.01465-09

39. Yazawa S, Yamazaki E, Saga Y, et al.: Evaluation of SARS-CoV-2 isolation in cell culture from nasal/nasopharyngeal swabs or saliva specimens of patients with COVID-19. Sci Rep. 2023, 13:8893. 10.1038/s41598-023-35915-w

40. Jeong HW, Kim SM, Kim HS, et al.: Viable SARS-CoV-2 in various specimens from COVID-19 patients. Clin Microbiol Infect. 2020, 26:1520–4. 10.1016/j.cmi.2020.07.020

